# Cohort profile: A Québec-based plasma donor biobank to study COVID-19 immunity (PlasCoV)

**DOI:** 10.1101/2022.11.09.22282156

**Authors:** Marc Germain, Antoine Lewin, Renée Bazin, Mélanie Dieudé, Josée Perreault, Amélie Boivin, Yves Grégoire, Christian Renaud

**Affiliations:** Medical Affairs and Innovation, Héma-Québec, Montréal and Québec, Canada

## Abstract

**Purpose:** Long-term humoral immunity to COVID-19 is not well understood owing to the continuous emergence of new variants of concern, the evolving vaccine- and infection-induced immunity, and the limited follow-up of previous studies. As the blood service in Québec (Canada), we established in April 2021 a COVID-19-focused biobank.

**Participants:** As of January 2022, included 86,229 plasma samples from 15,502 regular donors (age range=18-84 years, female %=49.7%), for an average of 5.6 donations per donor. Nearly two thirds (65.6%) of biobank donors made at least 2 donations, with many donors having provided samples pre- and post-vaccination (3061 [19.75%]) or pre- and post-infection (131 [0.85%]), thus allowing longitudinal studies on vaccine- and infection-induced immunity.

**Findings to date:** Comparative analysis of the immune response after the first and second dose of the BNT162b2 COVID-19 vaccine among SARS-CoV-2 naïve and previously infected individuals revealed that a single vaccine dose administered to previously infected individuals yields a maximal immune response. In contrast, SARS-CoV-2 naïve individuals required two vaccine doses to produce a maximal immune response. Furthermore, the results of a four-phase seroprevalence study indicate that the anti-N antibody response wanes quite rapidly, so that up to one third of previously infected donors were seronegative for anti-N.

**Future plans:** This plasma biobank from frequent and motivated donors, and the longitudinal nature of the biobank, will provide valuable insights into the anti-SARS-CoV-2 immune response and its persistence in time, and the effect of vaccination and of viral variants on the specificity of the antiSARS-CoV-2 immune response.

**Strengths and Limitations:** The herein described biobank has several strengths. To the best of our knowledge, this would be the largest biobank of plasma samples dedicated to COVID-19 research, with >80,000 samples from >15,000 donors and new samples continually being added until at least December 2022. Furthermore, the large subset of donors with ≥2 samples (65.6%) – along with the high frequency of donations in this subset (i.e., median: once every 29.0 days) – enables the conduct of longitudinal analyses on COVID-19 immunity. Another strength is that donors provided a broad consent, which allows researchers to recontact them for other projects (e.g., supplemental questionnaire). Lastly, the cost of establishing the biobank was minimized since the infrastructure and personnel required for sample collection were already in place at our blood collection sites. Given these strengths, our biobank may serve as a model for other blood operators and government partners who would be interested in reproducing our initiative elsewhere.

Certain limitations should nonetheless be considered when using our biobank samples. First, only plasma samples are available, such that the biobank cannot be used to study cell-based immunity. Researchers interested in studying cell-based immunity may want to contact BCQ19, which routinely collects peripheral blood mononuclear cells.[1] Second, despite the large sample, the plasma donor population is not fully representative of the overall Québec population, as expected since plasma donors are typically more representative of the healthy adult population. All exclusion criteria for plasma donations were also exclusion criteria for the biobank including immunodeficiencies, active infection, recent cancer among other chronic diseases. Third, the database associated with our biobank does not include information on disease severity, such as hospitalization or intensive care unit admission. Fourth, the database does not include information on socioeconomic status, such as income and education. However, the six-digit zip code can be used to generate a proxy index for socioeconomic status.

## Introduction

Despite remarkable progress, our understanding of the long-term, humoral immunity to COVID-19 is incomplete owing to a number of challenges. The emergence of new variants of concern, such as Omicron, continuously challenges prior assumptions and data on the effectiveness and persistence of COVID-19 immunity.[2,3] Furthermore, amid mass vaccination, disentangling infection and vaccine-induced humoral response to SARS-CoV-2 has become increasingly difficult to inform public health authorities.[4–6] Notably, the vast majority of published serological studies used a cross-sectional design, and hence do not provide individual-level data on COVID-19 immunity.[7] The more limited number of longitudinal studies that have been conducted had follow-up periods <12 months[8–19] and lacked long-term funding commitment. Clearly, new initiatives are needed to overcome these barriers.

Historically, biobanks have spurred research efforts on novel or rare diseases[20] and would likely help address the aforementioned challenges for COVID-19 research. They do so using highly standardized, quality-controlled processes to analyze a large number of biological specimens which are made available to the research community. In addition, biobanks are subject to regulatory oversight to protect donors’ rights. Virtually all biobanks collect a “broad” consent that allows for samples to be used for multiple research purposes.[21] For researchers, this practice (while controversial)[22] alleviates the burden associated with seeking consent.

Existing COVID-19-focused biobanks are suboptimal to longitudinally study COVID-19 immunity. The *Biobanque québécoise de la COVID-19* (BCQ19) is a biobanking initiative in Québec (Canada) that includes blood samples from individuals with a negative (controls) or positive COVID-19 PCR test result, with samples collected at several fixed time points for up to 24 months post-diagnosis (for non-hospitalized individuals) or post-hospitalization (for hospitalized individuals).[1] However, BCQ19 is limited in terms of sample size and population representativeness. Furthermore, the unavailability of samples prior to diagnosis or hospitalization hinders the study of the early immune response to COVID-19.

Blood services such as our institution (Héma-Québec – the only blood service operating in Québec) are ideally positioned to establish a biobank that would complement BCQ19 or other COVID-19-focused biobanks and better address researchers’ needs. Indeed, blood services routinely collect biological specimens from donors in the general population, thereby substantially alleviating the high cost normally associated with the establishment of a biobank. Blood services have been key partners in conducting COVID-19 research since the beginning of the pandemic;[23] involving them in biobanking efforts is therefore a sound approach to fuel additional research on COVID-19 immunity.

Here, we report on the establishment of a new biobank dedicated to COVID-19 research to inform public health in the province of Québec (Canada). The biobank consists of regular plasma donations collected throughout Québec, hence the name ‘PlasCoV’. Because plasma donors can donate every 6 days at our institution, the biobank includes a large number of repeat donors for whom longitudinal analyses are feasible.

### Cohort Description

The project was initiated on February 23, 2021. A pilot phase was launched in March 2021, and the project was expanded to all of our fixed collection centers in April 2021. At that time, the majority of the Québec general population was unvaccinated,[24] which enabled us to collect samples both pre- and post-vaccination obtained from many donors.

Ten fixed blood centers –all located in large or medium size (≥100.000 population) areas were designated to collect biobank-dedicated samples (**Table 1**). The resulting donor cohort is therefore broadly representative of plasma donors living in urban and suburban areas throughout Québec.

**Table 1.**
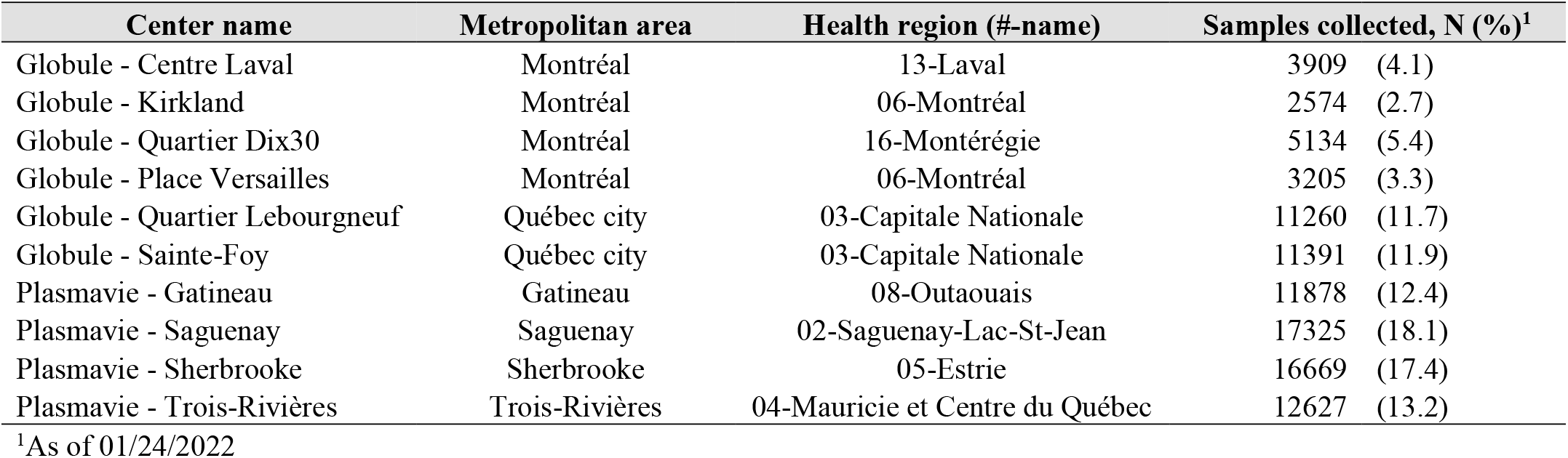
Number of samples collected by each blood center.

The project is entirely funded by the COVID-19 Immunity Task Force (CITF), a government-funded working group dedicated to advancing knowledge and research on COVID-19 in Canada and to inform public health authorities. CITF has provided a long-term funding commitment (2 years) for this biobank project, which will enable the conduct of long-term studies on infection- and vaccine-induced immunity to COVID-19.

The biobank includes samples from a population-based cohort of healthy voluntary, non-remunerated donors of plasma for fractionation who consented that their samples be used for research purposes. As of 01/24/2022, 15,502 out of 17,070 (90.8%) donors consented to have a small aliquot of their donations used for research on COVID-19. No donor recruitment campaign was specifically undertaken for this research initiative.

Donor characteristics were assessed as of 01/24/2022. Among consenting donors, the most common age group was 18-29 years (28.4%; **Table 2**). The six health regions where donors most often lived were Capitale-Nationale (18.1%), Estrie (14.7%), Saguenay-Lac-Saint-Jean (14.4%), Mauricie et Centre-du-Québec (11.4%), Outaouais (11.2%), and Montréal (9.8%). The cohort predominantly included donors who self-identified as white (93.6%). Relative to non-consenting donors, consenting donors tended to be younger (e.g., 18-29 years: 28.4% vs. 24.1% for the biobank and non-consenting donors respectively); and included slightly more female donors (i.e., 49.7% vs. 46.3%) and more residents of urban areas (e.g., Montreal-Laval region: 12.1% vs. 8.1%). Relative to the Québec general population, consenting donors had a similar age and sex distribution, but the densely populated region of Montreal-Laval was underrepresented (12.1% vs. 24.4%).

**Table 2.** Donor characteristics (as of 01/24/2022)

|  | Consenting donors<br>(N=15,502) |  | Non-consenting<br>donors<br>(N=1568) |  | General, adult<br>population<sup>1</sup> |
| --- | --- | --- | --- | --- | --- |
|  | N | (%) | N | (%) | (%) |
| **Sex<sup>2</sup>** |  |  |  |  |  |
| Female | 7709 | (49.7) | 831 | (46.3) | (50.2) |
| **Age (years)<sup>2,3</sup>** |  |  |  |  |  |
| 18-29 | 4399 | (28.4) | 433 | (24.1) | (17.1) |
| 30-39 | 2556 | (16.5) | 269 | (15) | (16.0) |
| 40-49 | 2287 | (14.8) | 251 | (14) | (16.1) |
| 50-59 | 2440 | (15.7) | 335 | (18.7) | (16.1) |
| 60-70 | 3178 | (20.5) | 408 | (22.7) | (18.3) |
| ≥71 | 642 | (4.1) | 100 | (5.6) | (16.4) |
| **Self-reported race/ethnicity<sup>2,4</sup>** |  |  |  |  |  |
| White | 14517 | (93.6) | 1667 | (92.8) | NA |
| Other | 985 | (6.4) | 129 | (7.2) | NA |
| **Health region (#-name)<sup>2,3</sup>** |  |  |  |  |  |
| 02-Saguenay-Lac-Saint-Jean | 2234 | (14.4) | 299 | (16.6) | (3.3) |
| 03-Capitale-Nationale | 2811 | (18.1) | 351 | (19.5) | (9.0) |
| 04-Mauricie et Centre-du-Québec | 1771 | (11.4) | 213 | (11.9) | (6.2) |
| 05-Estrie | 2282 | (14.7) | 277 | (15.4) | (5.8) |
| 06-Montréal | 1518 | (9.8) | 103 | (5.7) | (24.4) |
| 07-Outaouais | 1729 | (11.2) | 263 | (14.6) | (4.6) |
| 12-Chaudière-Appalaches | 744 | (4.8) | 100 | (5.6) | (5.0) |
| 13-Laval | 353 | (2.3) | 24 | (1.3) | (5.1) |
| 14-Lanaudière | 252 | (1.6) | 18 | (1) | (6.0) |
| 15-Laurentides | 333 | (2.1) | 22 | (1.2) | (7.4) |
| 16-Montérégie | 1259 | (8.1) | 108 | (6) | (16.6) |
| Regions without fixed centers<sup>4</sup> | 126 | (0.8) | <11 | (<0.70)<sup>5</sup> | (6.4) |
| Unknown or outside Québec | 90 | (0.6) | <11 | (<0.70)<sup>5</sup> | NA |
| **Number of vaccine doses received<sup>2,6</sup>** |  |  |  |  |  |
| 0 | 254 | (1.6) | NA | NA |  |
| 1 | 103 | (0.7) | NA | NA | (91.6) |
| 2 | 5275 | (34.0) | NA | NA | (88.7) |
| >2 | 9561 | (61.7) | NA | NA | (47.1) |
| Missing data<sup>7</sup> | 309 | (2.0) | NA | NA | NA |
| **Prior documented COVID-19 infection<sup>2,6,8</sup>** |  |  |  |  |  |
| Yes | 1662 | (10.7) | NA | NA | (5.6) |
| No | 13531 | (87.3) | NA | NA | (94.4) |
| Missing data<sup>7</sup> | 309 | (2.0) | NA | NA | NA |

**Abbreviation:** COVID-19 = coronavirus disease-19
**Notes:**
1. For the general population of Québec, various data sources were used to capture variables of interest 2. Demographics (i.e., sex, age, self-reported ethnicity, and health region) are reported as of 01-24-2022. Data on prior vaccination and documented infection are reported as of 01-24-2022. 3. For the general population, data come from the following: Ministère de la Santé et des Services sociaux du Québec. Estimations et projections de population par territoire sociosanitaire. Available from: https://publications.msss.gouv.qc.ca/msss/document-001617/. Accessed on August 10, 2022. 4. Includes (01-Bas-Saint-Laurent, 08-Abitibi-Témiscamingue, 09-Côte-Nord, 10-Nord-du-Québec, 11-Gaspésie-Îles-de-la-Madeleine, 17-Nunavik) 5. Exact number is lower than 11, and is thus not reported to preserve donor anonymity 6. For the general population, data come from the following: Institut national de santé publique du Québec. Données COVID-19 par âge et sexe - Évolution des cas. Available from: https://www.inspq.qc.ca/covid-19/donnees/age-sexe/evolution-cas. Accessed on 03-01-2022." 7. Missing data are due to mismatches between donor information recorded in our institution's database and that recorded in governmental databases (N=309), ) 8. Data for the general population are underestimated since the age groups used in publically available data (see footnote 7) exclude individuals aged 18-19 years.

The cohort includes 3061 donors with samples collected pre- and post-vaccination, thereby enabling the study of vaccine-induced immunity in a real-world population of vaccine-naïve individuals. The cohort also includes 10,551 donors with samples collected only post-vaccination. Finally, the cohort include 1326 donors with sample collected only pre-vaccination and 564 donors with no vaccination history or missing information. Another (more limited) subset consists of 131 donors with available data before and after a documented infection.

The vast majority (i.e., 96.4%) of donors had received ≥1 vaccine dose, and 95.7% had received ≥2 doses; only 1.6% were unvaccinated. In Québec, the second vaccine dose was delayed for up to 16 weeks to optimize population-level immunity amid concerns over limited vaccine supply. As a result, a large proportion of plasma donors (14.6%) received their second dose 85-112 days after their first dose, but some received it within the ?3 week gap recommended by the manufacturer (i.e., 1.9% within 4 weeks). This notable feature of our biobank may be used to study the impact of vaccine dose intervals on the immune response, as done by Tauzin et al.[25] (described in more details further down).

The following vaccines have been administered in Québec: ChAdOx1-S, BNT162b2, and mRNA-1273. Some individuals have received combinations of these vaccines, which was allowed by public health authorities to help fast-track the vaccination campaign.

As previously mentioned, a key feature of this biobank is the availability of multiple, longitudinal plasma samples and data collected for a large proportion of the cohort. As of 01/24/2022, the biobank had collected 86,229 samples from 15,502 donors, for an average of 5.6 donations per donor.

At least two donations are available for nearly two thirds of the cohort (i.e., 65.6%), thereby enabling the conduct of longitudinal analyses (**Figure 1**). Most of these repeat donors appeared to donate regularly, with a median gap between donations ranging from 51.0 days for those with 2 donations to 11.8 days for those with ≥15 donations (median=29 days among those with ≥2 donations; **Figure 1**).

**Figure 1.**
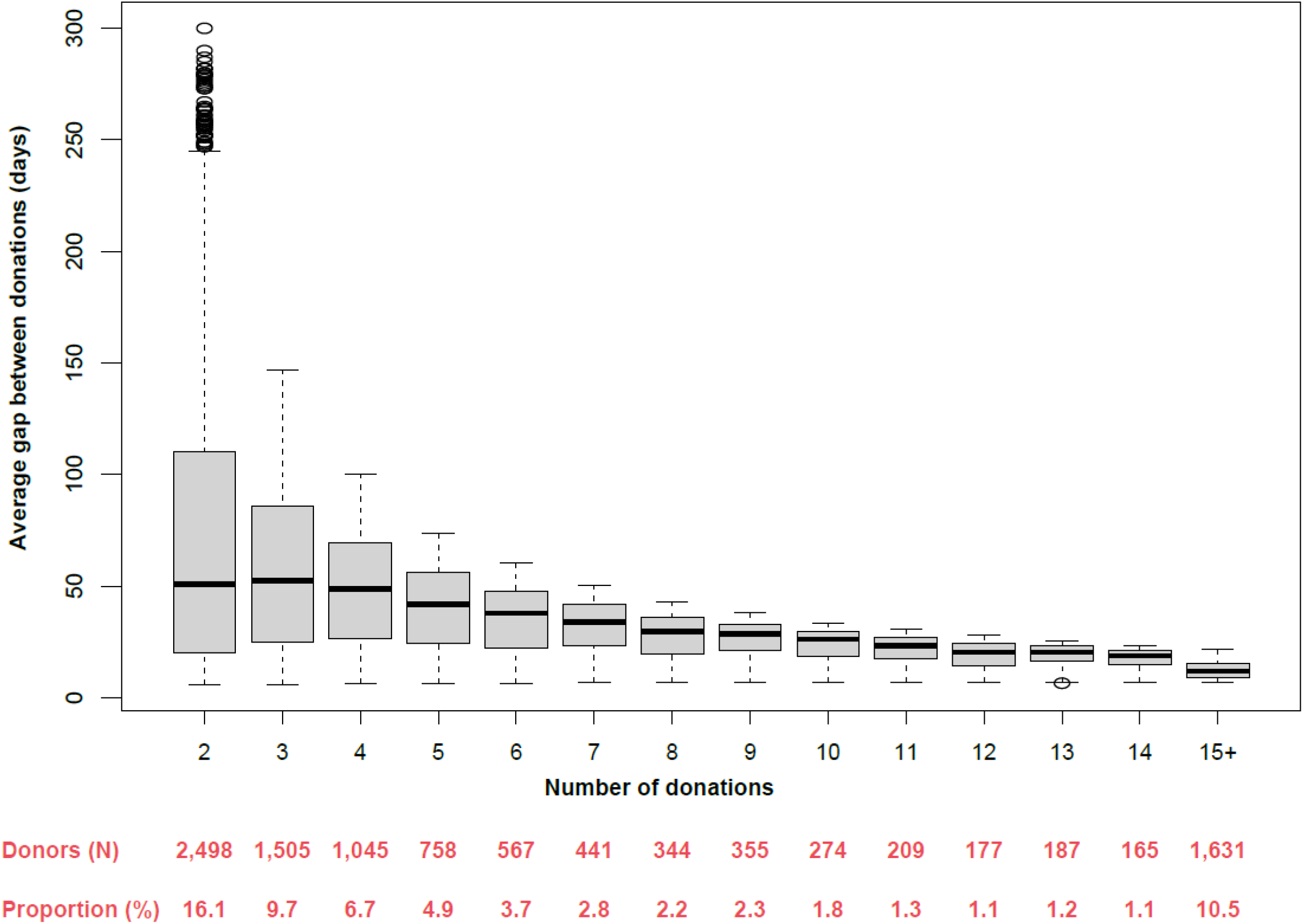
Average number of days between donations as function of the number of donations in PlasCoV.

Of note, loss to follow-up is not applicable to this cohort. Donors were free to donate plasma if and whenever they wanted: They did not consent to a strict protocol involving regular, scheduled visits at specific time points.

Several variables are collected as part of routine donor screening. These variables include demographic characteristics (e.g., age, sex) and clinical characteristics (e.g., blood type, recent blood-borne infections, diabetes; see **Table 3** for detailed list of measurements), which (except for blood type) are all self-reported by the donor. Other variables related to prior COVID-19 vaccination and infection were obtained by linking in-house donor data with the *Système d’information pour la protection des maladies infectieuses* (SI-PMI), a government vaccination registry Infection data was obtained through the government platform (Trajectoire de Santé Publique – TSP). This information was added to carry out a mandate given by the Ministère de la santé et des services sociaux for the seroprevalence study only. Of note, data on mortality and other comorbidities that are not relevant for plasma donation are not available in the dataset.

**Table 3.**
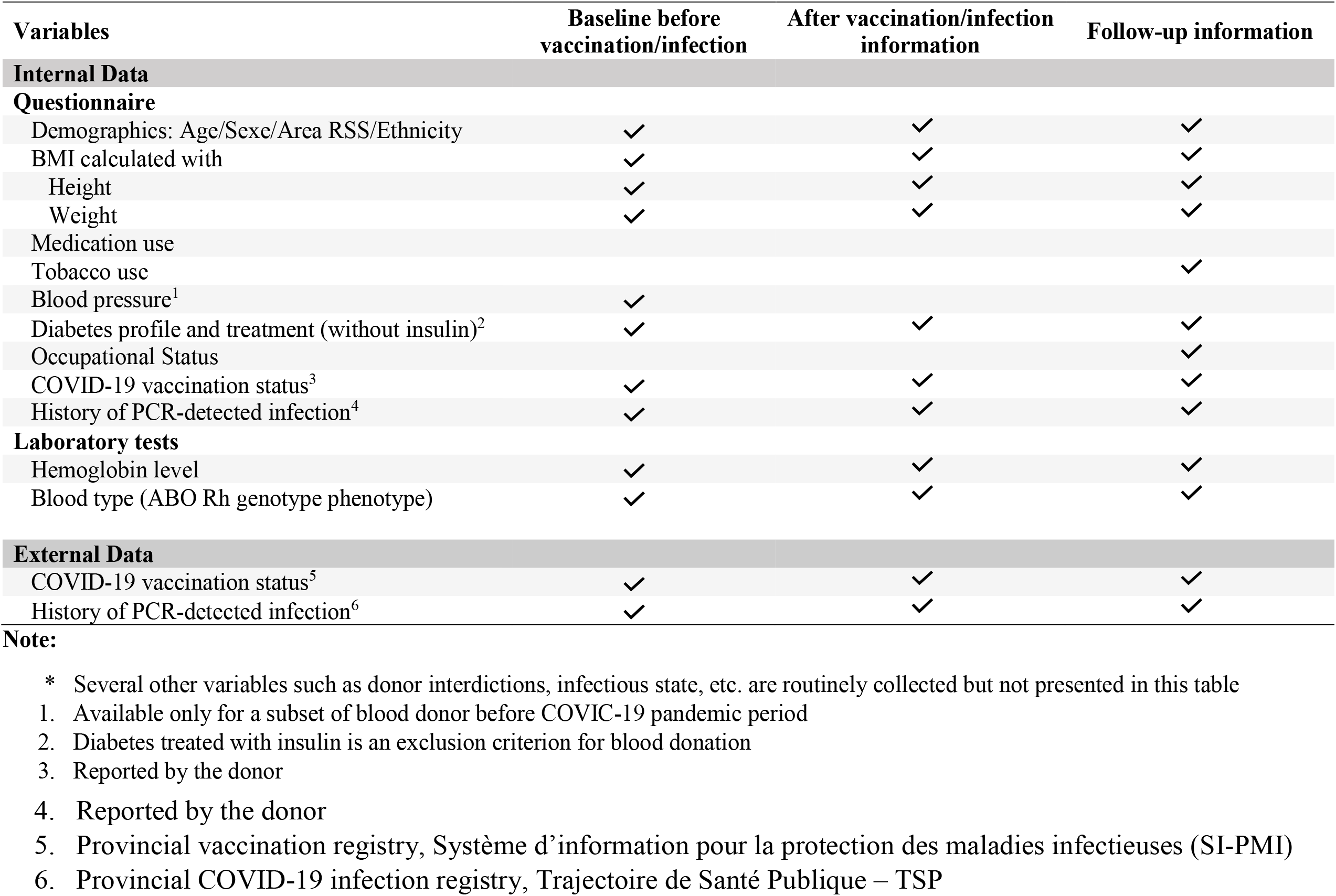
Summary of the principal* variables collected or derived from the cohort.

| Variables | Baseline before vaccination/infection | After vaccination/infection information | Follow-up information |
| --- | --- | --- | --- |
| **Internal Data** |  |  |  |
| **Questionnaire** |  |  |  |
| Demographics: Age/Sexe/Area RSS/Ethnicity | ✓ | ✓ | ✓ |
| BMI calculated with | ✓ | ✓ | ✓ |
| Height | ✓ | ✓ | ✓ |
| Weight | ✓ | ✓ | ✓ |
| Medication use |  |  |  |
| Tobacco use |  |  | ✓ |
| Blood pressure<sup>1</sup> | ✓ |  |  |
| Diabetes profile and treatment (without insulin)<sup>2</sup> | ✓ | ✓ | ✓ |
| Occupational Status |  |  | ✓ |
| COVID-19 vaccination status<sup>3</sup> | ✓ | ✓ | ✓ |
| History of PCR-detected infection<sup>4</sup> | ✓ | ✓ | ✓ |
| **Laboratory tests** |  |  |  |
| Hemoglobin level | ✓ | ✓ | ✓ |
| Blood type (ABO Rh genotype phenotype) | ✓ | ✓ | ✓ |
| **External Data** |  |  |  |
| COVID-19 vaccination status<sup>5</sup> | ✓ | ✓ | ✓ |
| History of PCR-detected infection<sup>6</sup> | ✓ | ✓ | ✓ |

**Note:**
\* Several other variables such as donor interdictions, infectious state, etc. are routinely collected but not presented in this table
1. Available only for a subset of blood donor before COVIC-19 pandemic period
2. Diabetes treated with insulin is an exclusion criterion for blood donation
3. Reported by the donor
4. Reported by the donor 5. Provincial vaccination registry, Système d'information pour la protection des maladies infectieuses (SI-PMI) 6. Provincial COVID-19 infection registry, Trajectoire de Santé Publique – TSP

### Patient and Public Involvement

Patients or the public were not involved in the design, or conduct, or reporting, or dissemination plans of our research

### Findings to Date

#### Longitudinal assessment of COVID-19 immunity

A study by Tauzin et al. used samples from our biobank to study the immune response to the mRNA vaccine BNT162b2 among SARS-CoV-2-naïve and previously infected (PI) individuals.[25] Specifically, anti-RBD titers, antibody binding, antibody-dependent cell-mediated cytotoxicity (ADCC), neutralization activity, and antibody avidity were assessed in the two groups after their first and second dose of vaccine. The impact of an extended interval (16 weeks vs 4 weeks) between the two doses of vaccine was also studied.

They observed that the levels of anti-RBD and anti-Spike antibodies as well as the capacity to recognize and neutralize different SARS-CoV-2 variants of PI individuals was increased after the first dose of vaccine and only minimally declined thereafter. Administration of the second dose to some PI individuals did not further increase the strength of the immune response compared to PI individuals vaccinated with only one dose.

In contrast, whereas the levels of SARS-CoV-2-specific antibodies as well as their functional activities increased in naïve individuals after the first dose of vaccine, the decline in antibody titers and functional activity was more pronounced compared to PI individuals. Administration of the second dose of vaccine permitted to reach much higher antibody levels than after their first dose. These levels were similar to those observed in PI individuals after their first or second dose of vaccine.

Tauzin et al. additionally investigated the impact of an extended vaccine interval among SARS-CoV-2-naïve individuals. With the exception of ADCC, all assays (antibody levels, variant recognition and neutralization) were consistent with a better immune response among individuals with an extended interval of ∼16 weeks compared with those with an interval of ∼4 weeks between the two doses.

#### Serosurvey

A publicly available report from our institution used biobank samples to estimate the seroprevalence of anti-SARS-CoV-2 antibodies in Québec from June 2021 – July 2021.[4] This was the third phase of a serosurvey initiated in May 2020. Phases 1 (May-June 2020) and 2 (January-March 2021) only included whole blood donors rather than plasma donors from the biobank, since the PlasCoV project had not yet been launched. Phase 3 assessed both anti-RBD and anti-N seroprevalence, since >80% of the population had received at least one vaccine dose at the time of the serosurvey.[24]

The anti-RBD seroprevalence was 89.61%, consistent with the widespread vaccination coverage during phase 3. However, the anti-N seroprevalence was only 6.43%, which was lower than the anti-RBD seroprevalence among unvaccinated blood donors included in phase 2 (i.e., 10.52%).

This unexpected result was likely driven by seroreversion, which occurs faster for anti-N than anti-S (or anti-RBD) antibodies.[26–28] Indeed, nearly 40% of PI donors in phase 3 tested seronegative for anti-N, likely because of seroreversion. This apparent rapid waning of anti-N antibodies was also observed in a separate cohort of 54 PI donors who donated convalescent plasma used in the CONCOR-1 clinical trial.[29] After >200 days of follow-up, anti-N seroreversion occurred in 33.3% of donors, whereas anti-RBD seroreversion occurred in only 11.1% of donors. Taken together, these results suggest anti-N seroprevalence may only be adequate to capture relatively recent infections, thereby questioning its usefulness in serosurveys.[6] At the time of writing this manuscript, a fourth seroprevalence study is ongoing with PlasCoV samples, in which we are comparing anti-N responses pre- and post-infection to estimate the incidence of recent infections in the context of the Omicron wave.

## Data Availability

All data produced in the present study are available upon reasonable request to the authors

## Collaboration

All researchers in Canada or elsewhere may apply to access the PlasCoV biobank. Researchers will be asked to fill a form with high-level information on their project’s objectives, methods, novelty, and relevance to the biobank’s objectives. They will also be asked to justify the sample size needed and any particular inclusion criteria, and obtain ethics approval for their project.

## Data Sharing Statement

Details on the PlasCoV biobank and the application process can be obtained through Héma-Québec’s website at https://www.hema-quebec.qc.ca/coronavirus/hema-quebec-en-contexte-de-pandemie/etude-plascov.en.html. Enquiries can be sent by e-mail to, or alternately to Dr. Marc Germain, the biobank director.

## Contributorship Statement

MG, AL, RB and CR conceived and designed the study. AL, AB, JP, and YG collected the data. AL and YG analyzed the data, with input from MG, RB, MD and CR. MG, AL, YG and RB drafted the manuscript and MD, AB, CR and JP critically revised it for important intellectual content. All authors approved the final version to be published.

## Ethics Approval

This study was approved by Héma-Québec Institutional review board. Individual informed consent was obtained from participants at the time of registration in the PlasCoV Biobank.

## Acknowledgments

Medical writing assistance was provided by Samuel Rochette, an employee of Héma-Québec.

